# Deficient and Altered Brain White Matter Development in Wolfram Syndrome

**DOI:** 10.64898/2026.05.27.26354240

**Authors:** Zhaolong Adrian Li, Olga Neyman, Jerrel Rutlin, Heather M. Lugar, Jonathan M. Koller, Joshua S. Shimony, Tamara Hershey

## Abstract

Wolfram syndrome (WFS) is characterized by youth-onset insulin-dependent diabetes and neurological deficits. Brain white matter deficiency has been reported, but its trajectory remains unclear. Applying diffusion basis spectrum imaging models longitudinally in 29 individuals with WFS (baseline ages, 5.2 to 25.8 years; maximum 7 visits) and 52 matched controls, we found that WFS is associated with microstructural alterations suggesting diminished axonal integrity, myelin content, and cellularity. These changes were present and stable early in the disease progression in visual and auditory-related regions, whereas abnormalities in the corpus callosum appeared later in adolescence and adulthood. Our results support developmental hypomyelination as a neurophenotype of WFS.

## 1 Introduction

Wolfram syndrome (WFS; OMIM #222300) is a rare, multisystem disorder that manifests as a combination of youth-onset insulin-dependent diabetes, optic atrophy, arginine vasopressin deficiency, hearing loss, gait and balance impairments, and neuropsychiatric concerns^1–4^. Estimates of WFS prevalence in the general population vary geographically from 1 in 160,000 in North America to 1 in 770,000 in the UK^3,5^. Amongst individuals with type 1 diabetes, prevalence is higher at 0.6% and is enriched in certain subgroups, including Lebanese (5.5%) and Han Chinese (1.1%) groups^6–8^, underscoring the importance of WFS research despite the disorder’s overall rarity.

The majority of individuals with WFS have autosomal recessive mutations in the *WFS1* gene (chromosome 4p16.1), which encodes wolframin, an endoplasmic reticulum (ER) transmembrane glycoprotein involved in regulating intracellular calcium flux and the unfolded protein response^9–13^. In WFS, aberrant or diminished production of wolframin disrupts ER and mitochondrial homeostasis, leading to cellular dysfunction and apoptosis. Such pathophysiology is well-characterized in the pancreas, where *WFS1* has been implicated in beta cell survival^14–16^. However, the multisystem presentation of WFS and the recognition of *WFS1*’s distributed expression throughout the body have prompted investigation into other organs^12^, including the brain, where atrophy and abnormalities in the brainstem, cerebellum, thalamus, optic nerve, and pituitary were amongst the first noted in case reports^3,17–22^.

Recently, systematic neuroimaging studies of WFS have identified reduced pontine, thalamic, and cerebellar volumes alongside widespread lower white matter integrity^12,23– 25^. In particular, findings involving white matter—myelin-ensheathed neurites connecting distal grey matter regions—have attracted interest given *WFS1*’s enriched expression in oligodendrocytes and astrocytes supporting myelination^12,26^. However, it remains unclear whether the observed lower white matter integrity and volume, primarily examined cross-sectionally to date, represented insufficient neurodevelopment or active neurodegeneration, which may have distinct mechanisms and require different treatment considerations. Such a question is optimally investigated using longitudinal data, especially because different white matter tracts mature at different rates^27,28^. Furthermore, advances in multicompartmental diffusion-weighted neuroimaging techniques, including diffusion basis spectrum imaging (DBSI)^29–32^, offer renewed potential to more precisely assess tissue microstructure and cellular environments than traditional volumetric and diffusion tensor imaging (DTI)-based approaches. Here, we characterized white matter microstructural development using DBSI in a longitudinal cohort of children and young adults with WFS.

## 2 Methods

### 2.1 Participants

Individuals with WFS were recruited into the Washington University Research Clinic natural history study from the Washington University Wolfram Syndrome International Registry or by self or physician referral. Eligibility for enrollment included confirmed *WFS1* mutations, self-awareness of diagnosis, baseline age under 30 years, and capability of travel to St. Louis, Missouri for yearly evaluations.

The control group consisted of healthy individuals as well as individuals with T1D who were otherwise healthy. Comparison between participants with WFS vs with T1D would help identify phenotypes specific to WFS unconfounded by diabetes complications. Participants with T1D were recruited from the Pediatric Diabetes Clinic at St. Louis Children’s Hospital. Exclusion criteria included diagnosis of psychiatric illness, significant neurological history unrelated to diabetes, premature birth (< 36 weeks of gestation) with complications, use of psychoactive medications, contraindications for neuroimaging, and physical conditions that would limit study participation. No control participant had neuropathy, retinopathy, or nephropathy.

Study procedures were approved by Washington University Human Research Protection Office. Youths gave informed assent (parents/guardians gave written informed consent). Adults gave written informed consent. Some of these recruitment details were included in previous reports^23,24^.

### 2.2 Neuroimaging

Multi-shell echo-planar diffusion-weighted magnetic resonance imaging was acquired between 2010 and 2017 on a Siemens MAGNETOM 3T Tim Trio system (Siemens Healthineers, Erlangen, Germany). Scans were collected up to 7 consecutive years for participants with WFS and 3 consecutive years for the control group. DTI and DBSI models were applied to assess tissue microstructure in 8 major association, commissural, and projection white matter tracts delineated by probabilistic tractography (**Figure 1**). DBSI partitions the water diffusion signal into anisotropic and isotropic compartments, which purportedly arise from different anatomical structures^29–32^. In contrast, the traditional DTI model makes no such distinction. Thus, DBSI has the potential to characterize tissue microstructure with greater specificity compared to DTI^29–32^. Like DTI, DBSI estimates voxel-wise axial diffusivity (AD; decreases with axonal injury), radial diffusivity (RD; increases with demyelination), and fractional anisotropy (FA; increases with overall white matter integrity and coherence), but these metrics are now unconfounded by isotropic diffusion^29–32^. DBSI also produces unique metrics, including fiber fraction (FF; increases with neurite density), nonrestricted fraction (NRF; increases with edema and tissue loss), and restricted fraction (RF; increases with cellularity)^29–32^. In white matter maturation, as myelination, refinement of axonal caliber, dendritic sprouting, and glial development happen, FA and restricted diffusion increase while RD and nonrestricted diffusion decrease^27,33^. See **Supplemental Methods** for further details on neuroimaging analysis.

**Figure 1.**
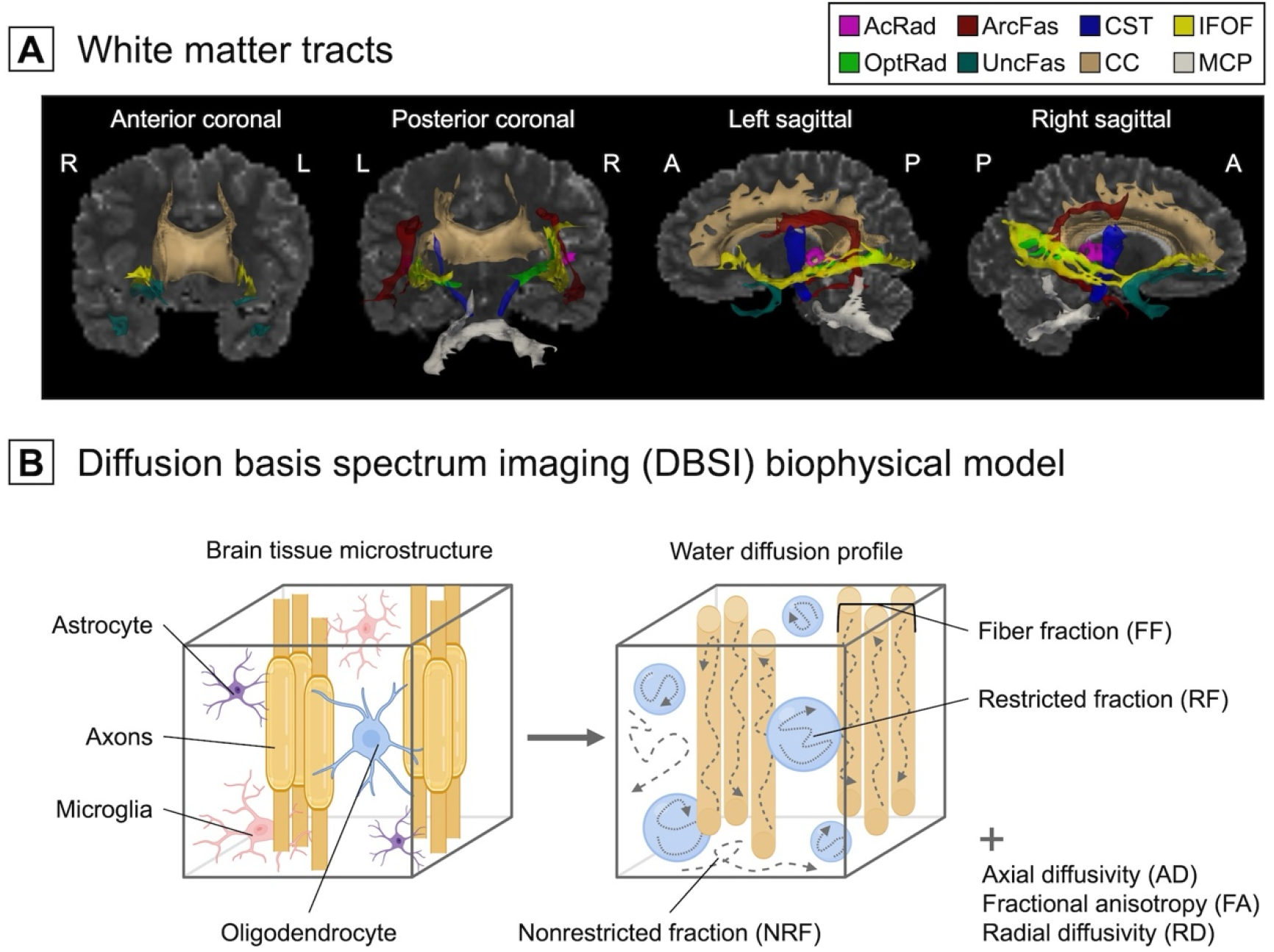
Characterization of Brain White Matter Microstructure. **(A)** Analyses focused on bilateral major association (arcuate fasciculus [ArcFas], inferior fronto-occipital fasciculus [IFOF], uncinate fasciculus [UncFas]), commissural (corpus callosum [CC]), and projection (acoustic radiations [AcRad], corticospinal tract [CST], optic radiations [OptRad], middle cerebellar peduncle [MCP]) tracts in the brain. A indicates anterior; L, left; P, posterior; R, right. **(B)** The diffusion basis spectrum imaging (DBSI) model separates the anisotropic and isotropic water diffusion signals, thereby modeling a range of neuroarchitectural properties. Figure was partly created with BioRender.

### 2.3 Statistical Analysis

Generalized additive mixed models with penalized splines (maximum basis dimension = 4) were used to compare longitudinal changes in microstructural metrics between groups, estimating both overall differences (ie, parametric term of group means) and trajectory differences (ie, factor-smooth term of group-by-age interactions). Models included sex and in-scanner head motion as covariates plus participant-level random intercepts and slopes. Significance was set at 2-sided false discovery rate (FDR)-corrected *P* value of .05. For significant group-by-age interactions, the first-order derivatives of nonlinear model fits and the accompanying 95% simultaneous CIs were estimated in 1/20^th^ of a year increments using the central finite difference method and 10,000 posterior simulations. Age ranges of significant development were defined as those with 95% CIs excluding 0 and were compared between WFS and control groups. Data were analyzed using R software version 4.3.1 (R Project for Statistical Computing).

## 3 Results

We analyzed 235 diffusion-weighted magnetic resonance imaging scans (**Supplemental Figure 1**) from 29 participants with pathogenic *WFS1* mutations (13 [45%] female; baseline ages, 5.2 to 25.8 years; maximum 7 scans) and 52 control participants (25 [48%] female; baseline ages, 6.0 to 26.2 years; maximum 3 scans). Within the control group, 24 participants had T1D. Groups were matched in terms of baseline age, sex, and in-scanner head motion (*P* ≥ .12) (**Table 1**).

**Table 1.**
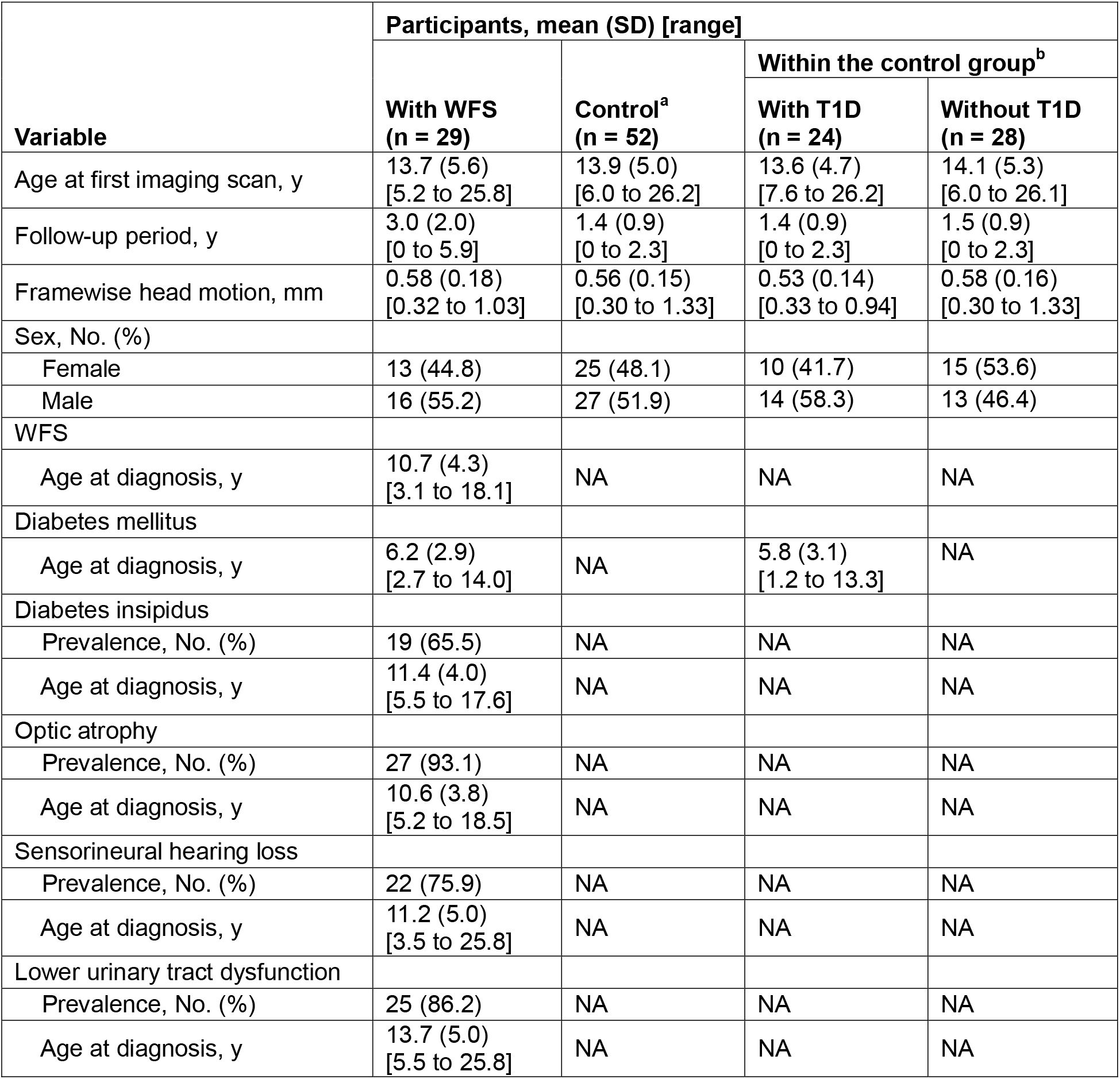
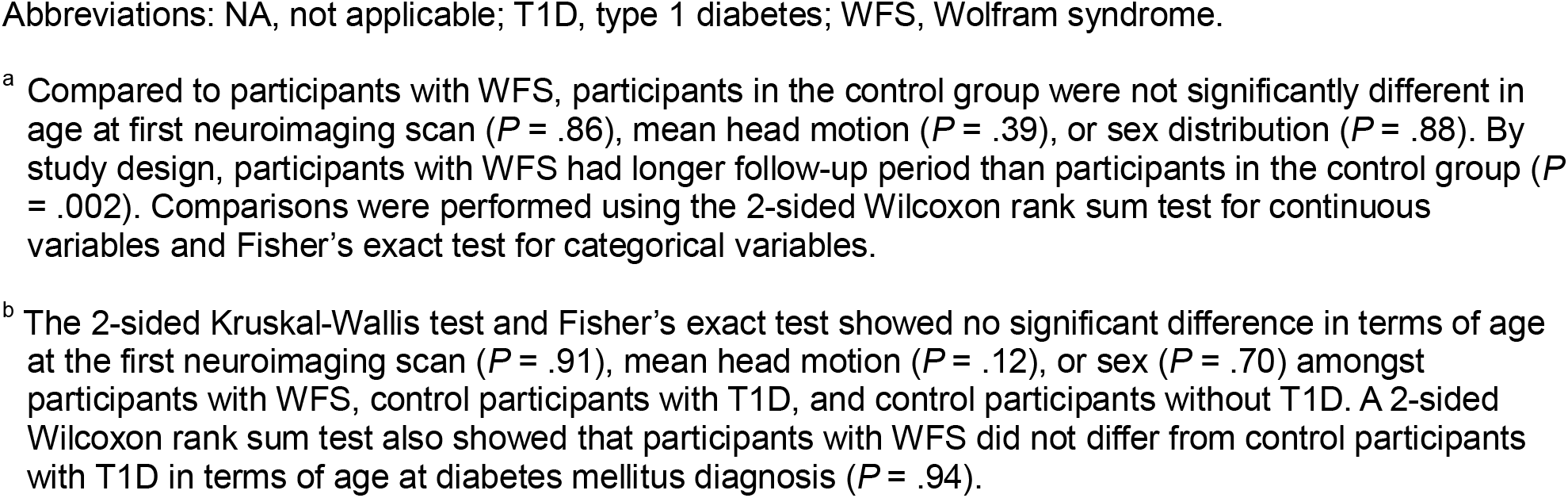
Participant Characteristics.

Overall group differences in white matter microstructure were widespread, with strongest effects in visual and auditory-related tracts including the optic radiations, inferior fronto-occipital fasciculus, and acoustic radiations, as well as regions involving the brainstem and cerebellum (ie, middle cerebellar peduncle) (**Figure 2; Supplemental Table 1**). These differences were driven by consistently lower fractional anisotropy, axial diffusivity, fiber fraction, and restricted fraction, as well as higher radial diffusivity and nonrestricted fraction in the WFS group compared to the control group across all ages studied. This overall pattern suggests widespread microstructural differences that could reflect lower axonal integrity, lower neurite density, lower cellularity, thinner myelin, and edema or tissue loss in WFS.

**Figure 2.**
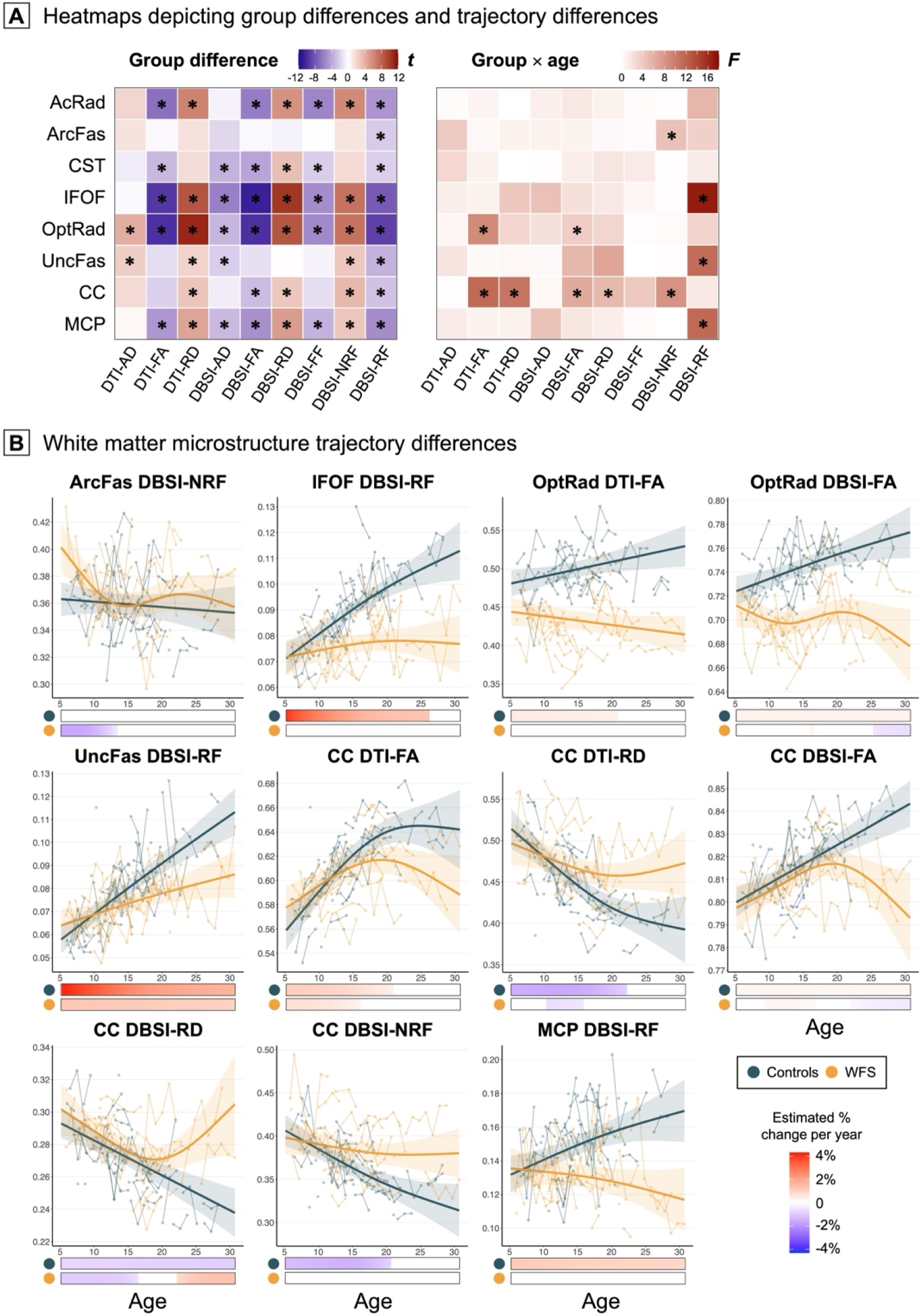
Overall and Developmental Trajectory Differences in White Matter Microstructure Between Wolfram Syndrome (WFS) and Control Groups. **(A)** Generalized additive mixed models estimated both overall differences and neurodevelopmental trajectory differences in brain microstructure between WFS and control groups. See **Supplemental Table 1** for model output. * indicates statistical significance, ie, two-tailed *P* ≤□.05 after false discovery rate correction. **(B)** For significant group-by-age interactions, nonlinear model fits were plotted as a function of age, flanked by shaded 95% confidence intervals (CIs) and overlaying individual participant-level trajectories. To characterize ages at which the group-level trajectories diverged, the first-order derivatives of model fits and the accompanying 95% simultaneous CIs were estimated in 1/20^th^ of a year increments using the central finite difference method and 10,000 posterior simulations. Age ranges of significant development, defined as those with 95% CIs excluding 0, are visualized in raster plots. AcRad indicates acoustic radiations; AD, axial diffusivity; ArcFas, arcuate fasciculus; CC, corpus callosum; CST, corticospinal tract; DBSI, diffusion basis spectrum imaging; DTI, diffusion tensor imaging; FA, fractional anisotropy; FF, fiber fraction; IFOF, inferior fronto-occipital fasciculus; MCP, middle cerebellar peduncle; NRF, nonrestricted fraction; OptRad, optic radiations; RD, radial diffusivity; RF, restricted fraction; UncFas, uncinate fasciculus.

Developmental trajectory differences were seen primarily with fractional anisotropy and radial diffusivity in the corpus callosum and restricted fraction in the inferior fronto-occipital fasciculus, uncinate fasciculus, and middle cerebellar peduncle, whereby participants with WFS exhibited levels and pacing of development comparable to control participants at early ages but demonstrated earlier plateauing and attenuated maturation thereafter. Most results remained when comparing the WFS group to the subset of control participants with T1D, indicating that observations were unlikely attributable to diabetes complications.

## 4 Discussion

In this study, we report longitudinal white matter microstructural development in the world’s largest and longest followed WFS cohort. The lower anisotropic water diffusion observed in WFS suggests diminished axonal integrity and myelin content^29–32^, and the lower restricted isotropic diffusion possibly reflects reduced neuronal/glial presence^29–32^, in line with the known enrichment of *WFS1* expression in oligodendrocytes and astrocytes^12^. As these diffusion metrics typically increase during normative white matter maturation^33^, our findings suggest that deficiencies in myelin and associated cells are present in WFS at an early age in most white matter tracts and persist into adulthood. Such interpretation is consistent with reports of axonal dystrophy with swelling in the pontocerebellar tract and optic radiations in post-mortem WFS samples^17,22^, as well as observations of aberrant neurite outgrowth in WFS patient-origin induced pluripotent stem cell (iPSC)-derived neural cells^34^.

However, the emergence of differences varied across white matter regions and age. For example, abnormalities in visual and auditory-related tracts were present and stable since the youngest age investigated, aligning with the early onset of sensory impairments in WFS^1–3^. However, it is possible that these differences emerged earlier than the youngest age scanned (5 years). In contrast, microstructural development in the corpus callosum appeared normal at early ages in WFS but diverged from normal trajectories in adolescence and adulthood. Some of the trajectories that diverged over age were with restricted fraction (in inferior fronto-occipital fasciculus; middle cerebellar peduncle and uncinate fasciculus), an index of intracellular water and model of cellularity, which could reflect diminished and stagnated developmental processes such as neurogenesis and glial maturation^33,35^. While we did not find changes in diffusion properties that would suggest neurodegeneration, we could not rule out such changes occurring at ages older than assessed here (age 26).

Our study has limitations. First, we lacked power to explore neurodevelopmental heterogeneity across different pathogenic variants of *WFS1*, which have been recently associated with differential disease severity^4^. Second, DTI and DBSI metrics are indirect measures of microstructure. Additionally, although insightful for group-level analyses, they demonstrate suboptimal within-individual reliability (as shown in **Figure 2**) and thus have limited utility as biomarkers for disease progression. Finally, the biological mechanisms behind our observed spatially patterned differences in developmental trajectories remain unclear.

Together, our results provide evidence for developmental hypomyelination as a neurophenotype of WFS. Animal models of WFS should ideally replicate this pattern so that molecular/cellular mechanisms could be investigated and early interventions might be developed to optimize neurodevelopment.

## Author Contributions

Mr. Li had full access to all of the data in the study and takes responsibility for the integrity of the data and the accuracy of the data analysis.

*Concept and design*: Li, Neyman, Shimony, Hershey

*Acquisition, analysis, or interpretation of data*: All authors.

*Drafting of the manuscript*: Li, Hershey

*Critical revision of the manuscript for important intellectual content*: Li, Shimony,

Hershey

*Statistical analysis*: Li

*Obtained funding*: Hershey

*Supervision*: Shimony, Hershey

## Conflict of Interest Disclosures

None reported.

## Funding/Support

This study was supported by the National Institutes of Health (NIH) *Eunice Kennedy Shriver* National Institute of Child Health and Human Development grants R01HD070855 (to Dr. Hershey) and U54HD087011 (to the Intellectual and Developmental Disabilities Research Center at Washington University), the NIH National Center for Research Resources grant UL1RR024992 (to Washington University Institute of Clinical and Translational Sciences), the NIH National Institute of Diabetes and Digestive and Kidney Diseases grant P30DK020579 (to the WashU Medicine Diabetes Research Center), The Snow Foundation, the American Diabetes Association, the George Decker and Julio V. Santiago Pediatric Diabetes Research Fund, the Mallinckrodt Institute of Radiology, and the McDonnell Center for Systems Neuroscience.

## Role of the Funder/Sponsor

The funding organizations had no role in the design and conduct of the study; collection, management, analysis, and interpretation of the data; preparation, review, or approval of the manuscript; and decision to submit the manuscript for publication. This manuscript is the result of funding in whole or in part by the National Institutes of Health (NIH) and so is subject to the NIH Public Access Policy. Through acceptance of this federal funding, NIH has been given a right to make this manuscript publicly available in PubMed Central upon the Official Date of Publication, as defined by NIH.

## Data Availability

The data that support the findings of this study are available on request from the corresponding author. The data are not publicly available due to privacy or ethical restrictions.

## Disclaimer

The content is solely the responsibility of the authors and does not necessarily represent the official views of the National Institutes of Health or other funders.

## Additional Contributions

The authors thank the other former and current Washington University Wolfram Study Group Members for advice and support in the greater research program and Mary Katherine Ray, PhD for help with figure creation, for which financial compensation was not received.

## Notes

### Competing Interest Statement

The authors have declared no competing interest.

### Author Declarations

Study procedures were approved by Washington University Human Research Protection Office (IRB ID # 201808060). Youths gave informed assent (parents/guardians gave written informed consent). Adults gave written informed consent.

