## Supplemental Methods, Supplemental Figure 1, and Supplemental Table 1 for "Deficient and Altered Brain White Matter Development in Wolfram Syndrome"

**Supplemental Methods.**
**Supplemental Figure 1.** Longitudinal Neuroimaging Data Included in Analyses
**Supplemental Table 1.** Generalized Additive Mixed Model Estimates
**Supplemental References.**

This supplemental material has been provided by the authors to give readers additional information about their work.

**Supplemental Methods.**

**Neuroimaging acquisition.** Participants with diabetes had their blood glucose levels measured before and after neuroimaging acquisition. Insulin doses were adjusted if pre-neuroimaging blood glucose was above 300 mg/dL, and food or juice was provided if levels were below 70 mg/dL. Diffusion-weighted magnetic resonance imaging was performed on a Siemens MAGNETOM 3T Tim Trio system (Siemens Healthineers, Erlangen, Germany) with the following parameters: echo-planar imaging sequence, 27 directions, b-values ranging from 0 to 1400 s/mm$^2$ (0, 0, 45, 112, 168, 224, 280, 336, 392, 448, 504, 560, 616, 672, 728, 784, 840, 896, 952, 1008, 1064, 1120, 1176, 1232, 1288, 1344, and 1400 s/mm$^2$), transverse acquisition, repetition time = 12300 ms, echo time = 108 ms, voxel resolution = 1.98 $\times$ 1.98 $\times$ 2 mm$^3$, acquisition time = 5:44 minutes; field map sequence, transverse acquisition, repetition time = 400 ms, echo time 1 = 4.92 ms, echo time 2 = 7.38 ms, voxel resolution = 4 $\times$ 4 $\times$ 4 mm$^3$, acquisition time = 0:54 minutes. Some of these details were included in previous reports[1,2].

**Neuroimaging analysis.** Diffusion-weighted images were skull-stripped and corrected for motion and eddy currents-induced distortions using the FMRIB Software Library (FSL)[3,4]. Images that passed visual quality inspection for motion artifacts, susceptibility artifacts, and extreme values were included in analyses (**Supplemental Figure 1**).

A standard diffusion tensor imaging (DTI) model was fit to diffusion-weighted images using FSL to derive measures of axial diffusivity (DTI-AD), fractional anisotropy (DTI-FA), and radial diffusivity (DTI-RD)[3]. Taking advantage of the multi-shell acquisition, we also fit a diffusion basis spectrum imaging (DBSI) model[5–7]. In DBSI, the total water diffusion signal is modeled by a linear combination of multiple discrete anisotropic tensors and a spectrum of isotropic tensors, whereas DTI models only a single "average" tensor. As DBSI's anisotropic and isotropic tensors are sensitive to different tissue microstructural properties, DBSI can characterize anatomy and pathology with greater granularity compared to DTI[5–7]. The DBSI equation is specified below:

$$S_k = \sum_{i=1}^{N_{Aniso}} f_i e^{-|\overrightarrow{b_k}|.\lambda_{\perp_i}} e^{-|\overrightarrow{b_k}|.(\lambda_{\parallel_i}-\lambda_{\perp_i}).\cos^2\Phi_{ik}} + \int_a^b f(D) e^{-|\overrightarrow{b_k}|D} dD \qquad (k = 1,2,3,\dots,k)$$

where $S_k$ and $b_k$ are the diffusion signal and the b-value of the $k^{\text{th}}$ diffusion gradient, $N_{Aniso}$ is the number of anisotropic tensors to be determined, $\Phi_{ik}$ is the angle between the $k^{\text{th}}$ diffusion gradient and the principal direction of the $i^{\text{th}}$ anisotropic tensor, $\lambda_{\parallel_i}$ and $\lambda_{\perp_i}$ are the axial and radial water diffusivities of the $i^{\text{th}}$ anisotropic tensor, $f_i$ is the signal intensity fraction for the $i^{\text{th}}$ anisotropic tensor, and $a$ and $b$ are low and high diffusivity limits of the isotropic diffusion spectrum $f(D)$.

The anisotropic metrics produced by DBSI included axial diffusivity (DBSI-AD; from $\lambda_{\parallel_i}$), fractional anisotropy (DBSI-FA), radial diffusivity (DBSI-RD; from $\lambda_{\perp_i}$), and fiber fraction (DBSI-FF; the sum of all anisotropic signal intensity fractions $f_i$). The isotropic measures produced by DBSI included nonrestricted fraction (DBSI-NRF; where the isotropic diffusion scale $D > 0.3$ μm²/ms, as in extracellular water) and restricted fraction (DBSI-RF; where $0 < D \leq 0.3$ μm²/ms, as in intracellular water)[5–7]. The biophysical interpretations of DBSI metrics are illustrated in **Figure 1** in the main text. Developmental neuroimaging studies have observed associations between white matter maturation and decreases in RD (purportedly reflecting thickening myelin)[8–13], decreases in AD (thought to reflect axonal refinement and crossing of fibers)[8–12], and increases in FA (growth in overall tissue integrity and coherence)[8–10,14–16]. Further, developmental increases in restricted isotropic/spherical diffusion, which RF models, have been observed and are thought to reflect growing numbers of neuronal and glial cell bodies and multi-directional cylindrical structures[14,17].

Probabilistic tractography (threshold at 1%) in FSL was used to delineate major white matter tracts on individual diffusion-weighted images[18,19], and the average DTI and DBSI metrics were extracted for each tract. Values that were more than 3 SD away from group mean across all timepoints were removed. The number of outliers was very limited: across 72 variables, mean (SD) of number of removed values = 1.2 (1.0), median = 1, minimum = 0, maximum = 4.

**Supplemental Figure 1.** Longitudinal Neuroimaging Data Included in Analyses

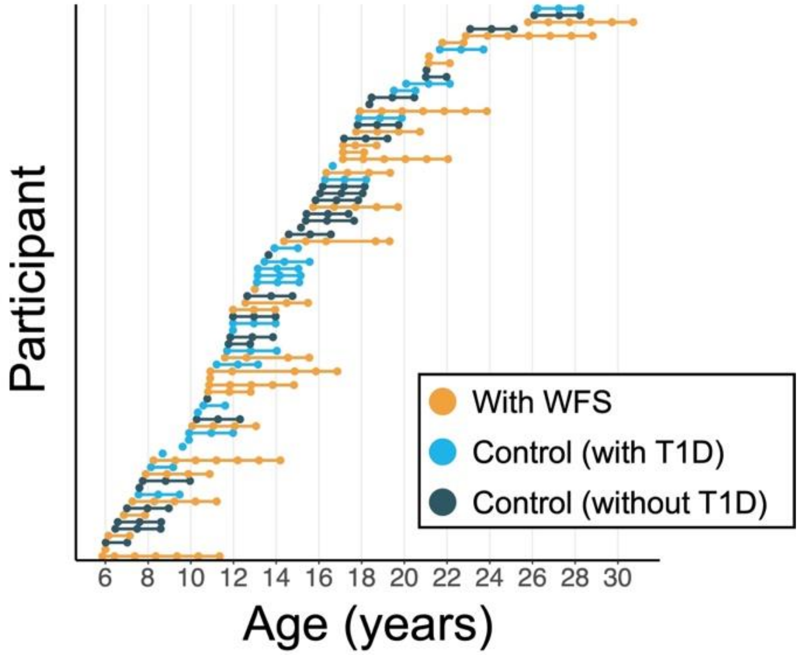


Scans that passed quality control are marked with dots, and data from the same participant are connected by a straight line. T1D indicates type 1 diabetes; WFS, Wolfram syndrome.

**Supplemental Table 1.** Generalized Additive Mixed Model Estimates

| White matter tract | Metric[a] | Group difference[b] | | | | Group × age interaction[b] | | |
|---|---|---|---|---|---|---|---|---|
| | | $\beta$ | $t$ | $P$ value | $P$ value (FDR)[c] | $F$ | $P$ value | $P$ value (FDR)[c] |
| Acoustic radiations | DTI-AD | 0.02 | 2.08 | .04 | .06 | 0.23 | .63 | .76 |
| | DTI-FA | -0.05 | -6.66 | < .001 | < .001 | 0.77 | .42 | .62 |
| | DTI-RD | 0.06 | 6.34 | < .001 | < .001 | 0.02 | .89 | .91 |
| | DBSI-AD | -0.004 | -0.57 | .57 | .63 | 1.22 | .27 | .50 |
| | DBSI-FA | -0.03 | -6.09 | < .001 | < .001 | 1.72 | .22 | .46 |
| | DBSI-RD | 0.04 | 5.55 | < .001 | < .001 | 1.03 | .32 | .54 |
| | DBSI-FF | -0.04 | -5.63 | < .001 | < .001 | 0.28 | .60 | .73 |
| | DBSI-NRF | 0.05 | 5.98 | < .001 | < .001 | 0.12 | .73 | .80 |
| | DBSI-RF | -0.01 | -4.87 | < .001 | < .001 | 5.49 | .02 | .09 |
| Arcuate fasciculus | DTI-AD | 0.01 | 1.41 | .16 | .19 | 3.91 | .05 | .16 |
| | DTI-FA | -0.002 | -0.24 | .81 | .83 | 0.52 | .47 | .64 |
| | DTI-RD | 0.01 | 1.50 | .13 | .17 | 0.88 | .44 | .62 |
| | DBSI-AD | -0.02 | -1.81 | .07 | .10 | 0.88 | .35 | .56 |
| | DBSI-FA | -0.001 | -0.32 | .75 | .79 | 0.89 | .31 | .54 |
| | DBSI-RD | -0.003 | -0.58 | .56 | .63 | 0.46 | .50 | .66 |
| | DBSI-FF | -0.0004 | -0.06 | .95 | .95 | 0.32 | .68 | .78 |
| | DBSI-NRF | 0.007 | 1.34 | .18 | .21 | 4.67 | .003 | .03 |
| | DBSI-RF | -0.005 | -2.32 | .02 | .03 | 3.55 | .06 | .18 |
| Cortico-spinal tract | DTI-AD | -0.02 | -0.86 | .39 | .45 | 2.98 | .09 | .24 |
| | DTI-FA | -0.04 | -3.14 | .002 | .004 | 1.03 | .38 | .59 |
| | DTI-RD | 0.02 | 1.76 | .08 | .11 | 0.01 | .93 | .95 |
| | DBSI-AD | -0.05 | -3.46 | < .001 | .002 | 0.09 | .76 | .82 |
| | DBSI-FA | -0.02 | -4.12 | < .001 | < .001 | 1.42 | .17 | .40 |
| | DBSI-RD | 0.02 | 3.24 | .001 | .003 | 1.91 | .08 | .24 |
| | DBSI-FF | -0.02 | -2.25 | .03 | .04 | 1.57 | .21 | .46 |
| | DBSI-NRF | 0.02 | 1.80 | .07 | .10 | 0.06 | .81 | .86 |
| | DBSI-RF | -0.004 | -2.13 | .03 | .05 | 1.43 | .23 | .47 |
| Inferior fronto-occipital fasciculus | DTI-AD | -0.002 | -0.27 | .79 | .82 | 1.25 | .27 | .50 |
| | DTI-FA | -0.06 | -9.49 | < .001 | < .001 | 1.07 | .29 | .52 |
| | DTI-RD | 0.07 | 8.73 | < .001 | < .001 | 4.22 | .04 | .15 |
| | DBSI-AD | -0.06 | -5.90 | < .001 | < .001 | 4.69 | .03 | .13 |
| | DBSI-FA | -0.05 | -10.68 | < .001 | < .001 | 2.33 | .05 | .16 |
| | DBSI-RD | 0.06 | 10.21 | < .001 | < .001 | 1.69 | .12 | .32 |
| | DBSI-FF | -0.03 | -5.26 | < .001 | < .001 | 0.64 | .42 | .62 |
| | DBSI-NRF | 0.04 | 7.17 | < .001 | < .001 | 0.15 | .70 | .78 |
| | DBSI-RF | -0.01 | -7.74 | < .001 | < .001 | 16.97 | < .001 | .007 |
| Optic radiations | DTI-AD | 0.04 | 4.01 | < .001 | < .001 | 0.64 | .42 | .62 |
| | DTI-FA | -0.07 | -9.91 | < .001 | < .001 | 8.89 | .003 | .03 |
| | DTI-RD | 0.11 | 10.94 | < .001 | < .001 | 3.13 | .02 | .09 |
| | DBSI-AD | -0.04 | -3.35 | < .001 | .002 | 2.37 | .13 | .32 |
| | DBSI-FA | -0.04 | -9.90 | < .001 | < .001 | 3.76 | .008 | .05 |
| | DBSI-RD | 0.06 | 8.67 | < .001 | < .001 | 3.12 | .02 | .09 |
| | DBSI-FF | -0.03 | 5.41 | < .001 | < .001 | 0.21 | .65 | .76 |

| | | | | | | | | |
|---|---|---|---|---|---|---|---|---|
| | DBSI-NRF | 0.05 | 7.16 | < .001 | < .001 | 0.34 | .56 | .71 |
| | DBSI-RF | -0.02 | -9.06 | < .001 | < .001 | 3.23 | .02 | .09 |
| Uncinate fasciculus | DTI-AD | 0.02 | 2.45 | .02 | .03 | 2.06 | .15 | .38 |
| | DTI-FA | -0.01 | -1.56 | .12 | .16 | 0.47 | .49 | .66 |
| | DTI-RD | 0.02 | 2.19 | .03 | .05 | 0.59 | .44 | .62 |
| | DBSI-AD | -0.02 | -2.25 | .03 | .04 | 0.41 | .52 | .67 |
| | DBSI-FA | -0.006 | -1.42 | .16 | .19 | 5.43 | .02 | .09 |
| | DBSI-RD | 0.0005 | 0.08 | .94 | .95 | 6.81 | .01 | .06 |
| | DBSI-FF | -0.003 | -0.51 | .61 | .67 | 0.002 | .96 | .96 |
| | DBSI-NRF | 0.02 | 2.84 | .005 | .009 | 0.14 | .71 | .78 |
| | DBSI-RF | -0.007 | -3.46 | < .001 | .002 | 11.35 | < .001 | .02 |
| Corpus callosum | DTI-AD | 0.02 | 1.82 | .07 | .10 | 0.14 | .70 | .78 |
| | DTI-FA | -0.01 | -2.04 | .04 | .06 | 11.51 | < .001 | .02 |
| | DTI-RD | 0.02 | 2.97 | .004 | .006 | 10.61 | .001 | .02 |
| | DBSI-AD | -0.006 | -0.83 | .41 | .47 | 0.57 | .45 | .62 |
| | DBSI-FA | -0.008 | -3.03 | .003 | .006 | 6.44 | .001 | .02 |
| | DBSI-RD | 0.01 | 2.80 | .006 | .01 | 5.37 | .004 | .03 |
| | DBSI-FF | -0.007 | -1.49 | .14 | .17 | 4.14 | .04 | .15 |
| | DBSI-NRF | 0.02 | 3.53 | < .001 | .001 | 8.26 | .005 | .03 |
| | DBSI-RF | -0.005 | -2.76 | .006 | .01 | 1.64 | .20 | .45 |
| Middle cerebellar peduncle | DTI-AD | 0.007 | 0.39 | .70 | .75 | 1.48 | .23 | .47 |
| | DTI-FA | -0.06 | -4.78 | < .001 | < .001 | 0.87 | .35 | .56 |
| | DTI-RD | 0.06 | 4.59 | < .001 | < .001 | .98 | .32 | .54 |
| | DBSI-AD | -0.04 | -3.14 | .002 | .004 | 4.60 | .03 | .13 |
| | DBSI-FA | -0.04 | -5.07 | < .001 | < .001 | 1.88 | .17 | .40 |
| | DBSI-RD | 0.04 | 5.03 | < .001 | < .001 | 1.31 | .25 | .49 |
| | DBSI-FF | -0.04 | -3.45 | < .001 | .002 | 0.28 | .59 | .73 |
| | DBSI-NRF | 0.04 | 2.87 | .005 | .008 | 0.04 | .85 | .89 |
| | DBSI-RF | -0.02 | -5.26 | < .001 | < .001 | 11.48 | < .001 | .02 |

Abbreviations: AD, axial diffusivity; DBSI, diffusion basis spectrum imaging; DTI, diffusion tensor imaging; FA, fractional anisotropy; FDR, false discovery rate; FF, fiber fraction; NRF, nonrestricted fraction; RD, radial diffusivity; RF, restricted fraction.

[a] See **Supplemental Methods** for details on neuroimaging acquisition and the derivation of these metrics.

[b] Generalized additive mixed models with penalized splines (maximum basis dimension 4) included a parametric term of group (ie, participants in the Wolfram syndrome group vs. control group), a factor-smooth term of group-by-age interaction, covariates of sex and in-scanner head motion, and participant-level random intercepts and slopes.

[c] Cells with two-tailed $P \leq .05$ after false discovery rate correction are highlighted.

**Supplemental References.**
